# Exploring the relationship between stroke lesion characteristics and sleep in chronic stroke survivors

**DOI:** 10.1101/2025.09.22.25336322

**Authors:** Anna á V. Guttesen, Matthew Weightman, Barbara Robinson, Katrijn B. Schruers, Pei-Ling Wong, Hanna E. Willis, Holly Bridge, Charlotte J. Stagg, Ioana-Florentina Grigoras, Oana M. Puicar, Heidi Johansen-Berg, Melanie K. Fleming

## Abstract

Sleep is often disrupted after stroke. However, little is known about how stroke lesion extent or location influences sleep, particularly at the chronic stage of recovery. In this pragmatic study, we aimed to explore whether lesion characteristics could explain sleep variability in chronic stroke survivors. We analysed previously collected structural brain images (Magnetic Resonance Imaging) from 38 stroke participants (11 female, mean (SD) age 64 (12), mean (SD) time since stroke 95 (66) months) and sleep data (questionnaires (N=38, actigraphy (N=37), and electrophysiology (N=18)) which were collected in their home environment. Neither lesion volume nor lesion overlap with regions of interest (brainstem, basal ganglia, amygdala, hippocampus, thalamus) significantly predicted interindividual variability in subjective or objective sleep measures. However, a data-driven approach revealed clusters of voxels disconnected by the stroke lesions were linked to lower spindle density and amplitude (threshold free cluster enhancement p < 0.050). Overall, these results provide preliminary insights that lesion induced brain disconnection, rather than the extent and overlap of the lesion with grey matter regions, may be more informative when explaining sleep variability. However, larger prospective studies are needed to fully understand the effect of stroke lesions on sleep.

## Introduction

Sleep is frequently reported as disrupted after stroke (Baylan et al., 2020; Gottlieb et al., 2019). This disruption includes altered sleep-wake patterns, fragmented sleep, and difficulties initiating or maintaining sleep (Baillieul et al., 2023). Systematic reviews report that sleep disorders are more common after stroke than for people without stroke (Gottlieb et al., 2019), showing prevalence of insomnia as up to 60% (Baylan et al., 2020). Sleep problems are associated with poorer recovery outcomes and participation (Fulk et al., 2020), but little is known about why sleep is disrupted after stroke. Sleep disturbance may be a consequence of multiple modifiable factors, including disruptive hospital environments, concomitant conditions such as low mood or pain, and distortion of sleep-wake cycles with reduced sleep pressure resulting from altered behaviours or routines (Cai et al., 2021; Wesselius et al., 2018). However, the impact of the lesion itself on the neuroanatomical circuitry involved in the regulation of the sleep-wake cycle, and hence, sleep outcomes, is not yet well understood.

There are limited studies examining whether lesion location or lesion extent relates to sleep at either the acute or the sub-acute stage of recovery after stroke. Liu et al. (2023) found that patients with posterior circulation stroke had significantly less stage 3 (slow wave) non-rapid eye movement (NREM) sleep and more wake after sleep onset (WASO) at the acute stage of stroke compared to patients with anterior circulation infarcts. However, participants were included in the study because they had a sleep assessment due to sleep problems. Therefore, whilst this study is useful to provide some understanding of how sleep problems might differ based on lesion location, the sample is likely biased towards people with clinically significant sleep disruption. Mekky et al. (2023) found that patients with stroke infarcts to brainstem had significantly more REM sleep and longer REM latency compared to patients with other subcortical and cortical infarcts. Gottlieb et al. (2020) found lower amygdala volume for stroke survivors with low sleep efficiency (<80%) compared to those with “optimal” sleep efficiency (≥80%), as well as reduced volume of the thalamus, amygdala and pallidum for those with long sleep duration compared to healthy controls. However, this study deliberately excluded any participants with lesions in their regions of interest (thalami, hippocampi, amygdala, basal ganglia, and brainstem), which limits the direct applicability of these results to understanding lesion-related sleep alterations. Importantly, these studies investigated sleep at the acute and sub-acute stage of recovery, but sleep problems have also been identified at the chronic stage (Fleming et al., 2021; Hasan et al., 2021). Thus, there is currently a considerable gap in our understanding of the impact of stroke lesions on sleep continuity and architecture at the chronic stage of recovery.

Even less is understood about how stroke lesions impact the underlying brain oscillation characteristics of sleep. In particular, the potential impact on slow oscillations and sleep spindles warrants attention given their relevance to learning and neuroplasticity which could influence recovery and rehabilitation (Diekelmann & Born, 2010; Duss et al., 2017). In an experimental (rodent) model of stroke, Kim et al. (2022) demonstrated that coupling of slow oscillations and spindles was impaired acutely, but normalised with recovery. One human study found that larger lesion volume was associated with reduced amplitude of sleep spindles, but they were unable to investigate whether the location of the lesion was a factor due to the limited sample size (Gottselig et al., 2002). It is important to probe the impact of stroke on neural oscillations during sleep to better understand how sleep could impact on stroke recovery. We have previously demonstrated that sleep disruption is associated with worse motor outcomes during rehabilitation after stroke and brain injury (Fleming et al., 2020), and hypothesise that this may be, at least in part, due to the disruption to neuroplasticity processes required to consolidate learning during rehabilitation.

The aim of the present study was therefore to test whether lesion characteristics explain variability in sleep outcomes at the chronic stage of stroke recovery. In the longer term, determining which factors are associated with poor sleep after stroke could help clinicians to identify patients at risk of long-term sleep complications, with a view to early intervention. To begin to address this aim, we analysed previously collected structural brain images of stroke survivors at the chronic stage of recovery, combined with a mixture of previously collected and new data on participants’ sleep in their home environment (using questionnaires, actigraphy, and low-density electroencephalography). This study serves as a preliminary exploration to identify potential candidate brain regions to investigate in future prospective studies. We hypothesised that lesions overlapping areas involved in regulation of the sleep-wake cycle (e.g. thalamus, hippocampus, striatum, brainstem) would be associated with worse sleep measures.

## Methods

This pragmatic study made use of an existing dataset of magnetic resonance imaging (MRI) scans obtained at the University of Oxford across multiple studies (ethics approval references R74800, R59721, 14/LO/0020, R60132). Inclusion criteria were: stroke survivors for whom a T1-weighted structural MRI scan was available, aged 18 years or over, discharged from inpatient rehabilitation, and living in the United Kingdom. Exclusion criteria were: current shift workers, and people with neurological or psychiatric conditions other than stroke that the researcher deemed could affect participation (e.g. Parkinson’s Disease).

Existing sleep data were available for some participants who had participated in other studies run by the research team. Additional data were sought by contacting participants who had agreed to be contacted about other research opportunities. Following written informed consent, participants were sent questionnaires to complete (either online through Jisc or in paper format depending on their preference). They were also provided with an actigraphy monitor and a low-density electroencephalography headband to wear at home. The study was approved by the Central University Research Ethics Committee (reference R85306) and the aims, methods and parameters of interest were preregistered prior to any data analysis using the Open Science Framework (https://osf.io/khqn8).

### Participants

Participant characteristics are shown in table 1. MRI scans were available for 57 stroke survivors. Sleep data from previous studies in our centre were available for 14 of the participants. The remaining 43 participants were invited to provide sleep data and 30 participants consented to do so (see figure S1 for description of data availability). The time between data collection of the MRI scans and sleep measures was 22 (± 26) months. Participants were compensated for their time via £30 online shopping voucher or bank transfer.

**Table 1.** Participant characteristics.

|  | Mean (SD) or count |
| --- | --- |
| Age, years | 64 (12) |
| Male/female | 27/11 |
| Time since stroke, months | 95 (66) |
| Lesioned left/right hemisphere | 11/27 |
| Estimated Body Mass Index (N=25) | 27 (4) |
| Type of stroke (N=26) |  |
| *Ischaemic* | 16 |
| *Haemorrhagic* | 4 |
| *Unknown* | 6 |
| Ethnic Group |  |
| *White British/Irish/European* | 28 |
| *Black/African/Caribbean/Black British* | 1 |
| *Other ethnic group* | 2 |
| *Unknown* | 7 |
| Medications (N=31) |  |
| *SSRI/SNRI* | 5 |
| *TCA* | 2 |
| *Anticonvulsants* | 5 |
| *Antihistamine* | 1 |

Values are mean +/- standard deviation or count. SSRI: selective serotonin reuptake inhibitor; SNRI: serotonin norepinephrine reuptake inhibitor; TCA: tricyclic antidepressant. Anticonvulsants include Sodium Valproate, Pregabalin, Gabapentin, Levetiracetam.

#### Magnetic Resonance Imaging (MRI)

MRI data were acquired on a 3 Tesla Siemens Magnetom Prisma MRI scanner (Siemens Healthcare AG) using either a 32-channel or 64-channel head coil. A whole brain anatomical T1-weighted magnetization prepared rapid gradient echo (MPRAGE) image was collected with voxel-size=1×1×1mm, flip angle=8°, TR=1900 ms. Head movement was minimised by use of foam padding.

#### Questionnaires

Participants were asked to complete a demographic questionnaire, including information regarding their age, sex, approximate weight and height (to approximate body mass index; BMI), medications, stroke details (e.g. date, type of stroke if known). We also used the modified Rankin Scale score as a measure of post-stroke disability.

To assess sleep and sleep disorders we used the Sleep Condition Indicator (SCI), Pittsburgh Sleep Quality Index (PSQI), STOP BANG (to assess potential risk for Sleep Apnoea), Sleep Disorders Screening Questionnaire (SDSQ) and the Epworth Sleepiness Scale (ESS). To assess mood, we used the Patient Health Questionnaire (PHQ-9) and the Generalised Anxiety Disorder Questionnaire (GAD-7). To assess physical activity, we used the International physical activity questionnaire short form (IPAQ-SF) which is measured in MET (MET or MET-minutes are multiples of the resting metabolic rate and calculated by weighting each activity by its energetic requirements) and time spent sitting.

#### Actigraphy

Participants were asked to wear a waterproof actigraphy monitor on their less-affected wrist (Motionwatch8, CamNtech Ltd., UK) for 1 week. The Motionwatch 8 contains an accelerometer and motion is converted into an activity count, using 30 s epochs. To facilitate data analysis, participants were asked to complete a short sleep diary each morning to indicate the approximate time they tried to sleep the previous night, and the time that they woke up (prior to getting up in the morning) while wearing the monitor. They were also asked to indicate via button press when they switched off the lights to go to sleep. A combination of the information provided was used to determine the bedtime period.

#### Low-density Electroencephalography (EEG)

Participants were asked to wear a low-density EEG headband (Dreemband II and III, Dreem Inc, France) for at least three nights to record their sleep EEG at home. This EEG headband is commercially available and was developed and validated with the aim of providing an accessible means of monitoring sleep outside of a sleep laboratory (Arnal et al., 2020). Velcro size-adjusters can be used to adjust the circumference of the headband as needed and participants were supplied with an elastic headband to place over the device if needed to ensure it was secure. The Dreemband uses 2 dry, silicone frontal electrodes at F7 and F8, two occipital electrodes at O1 and O2, and a ground at FP2 according to the international 10-20 system. Data are transferred to the server using Bluetooth and were downloaded by researchers in EDF format. Automated sleep scoring by Dreem was also exported as a hypnogram text file (Arnal et al., 2020).

#### Sample size

The sample size for this study was dependent on how many previous participants with a T1-weighted scan we were able to obtain sleep data for. We aimed to include a minimum of 30 participants, which would enable us to detect a correlation of r = 0.5 (80% power and alpha = 0.05) between degree of lesion overlap with an individual region of interest (ROI) and a sleep measure of r=0.5. However, once we obtained and began analysing the data it became clear that the planned correlation analysis would not be suitable, given the high number of participants with a lesion-ROI overlap of 0. Thus, we binarised the overlap maps and characterised overlap as yes/no for each ROI.

### Analyses

#### Magnetic Resonance Imaging (MRI)

##### MRI preprocessing

All preprocessing steps were carried out in FSL (Jenkinson et al., 2012). Stroke lesions were manually segmented using T1-weighted MRI images in *fsleyes*, and subsequently smoothed with 2mm full width half maximum (FWHM; *fslmaths*, lower threshold=35). Brain extraction was performed using the optiBET tool (Lutkenhoff et al., 2014). Finally, non-linear registration and transformation to standard MNI152 1mm space was conducted using the inverted lesion mask (*fnirt* and *applywarp*). Lesion heatmaps in standard space were generated using *fslmaths*.

##### MRI regions of interest (ROIs)

The Harvard-Oxford probabilistic atlases were transformed from standard to native space using the non-linear transform with a probability threshold of 60%. A priori regions of interest included brainstem, basal ganglia (nucleus accumbens, caudate and pallidum), amygdala, hippocampi and thalami (all regions bilateral). Lesion volume and lesion overlap with regions of interest were calculated using *fslstats.* Lesion overlaps with regions of interest were calculated separately for each hemisphere, and subsequently collapsed to get one number per each region of interest.

##### Structural connectivity networks

To assess structural disconnections, we used the BCBtoolkit (v4.1.0) which is described in detail in Foulon et al. (2018). In short, this approach tracks fibres in diffusion weighted imaging datasets from 10 healthy controls and registers each patient’s lesions from MNI152 space to each control’s native space, so that they can be used as seed for tractography in Trackvis (Wang et al., 2007). Tractographies from the lesions are transformed in visitation maps (De Schotten et al., 2011), binarised and transformed to MNI space, and finally a percentage overlap map is created by summing at each point in MNI space the normalised visitation map of each healthy subject. This will result in one disconnectome map for each patient. In the resulting disconnectome map, the value in each voxel provides the probability of disconnection from 50 to 100% for a given lesion (Thiebaut de Schotten et al., 2015).

#### Actigraphy

Actigraphy data were extracted using the custom software, Motionware (Camntech Ltd, UK) with a threshold of 20 (high sensitivity). The median value of the 7 nights was obtained for sleep measures: assumed sleep time (i.e. the time spent in bed with the intention of sleeping, minutes), sleep efficiency (SE, i.e., estimated actual sleep time as a percentage of assumed sleep time), wake after sleep onset (WASO; minutes), number of awakenings (wake bouts), and the sleep fragmentation index (the sum of the total time categorised as mobile, expressed as a percentage of the assumed sleep, and the number of immobile bouts which were <1 min in length, expressed as a percentage of the total immobile bouts). Note, SE was not determined from sleep onset (an oversight in the preregistration), but from assumed sleep time and actual sleep time, thus ensuring compatibility with the literature. We chose to focus predominantly on measures of sleep duration and continuity as these have previously been found to relate to volumetric brain changes post-stroke (Gottlieb et al., 2020) and to be different for stroke survivors compared to controls (Fleming et al., 2021).

#### Electroencephalography (EEG)

##### Sleep staging

The raw data were visually inspected and sleep stages (30 s epochs) were manually rescored (revised from Dreem automated scoring) by trained sleep scorers according to the AASM (Iber, 2007), with highly noisy epochs scored as artifacts (mean ± SD percentage: 6.00 ± 6.13). The rescored hypnograms were used for further analyses. Data from 3 participants were deemed too noisy for sleep staging (final N=17), and the number of sessions per participant varied from 1-4 (mean ± SD: 2.06 ± 0.90). For the main analyses, median values were calculated across the included nights.

##### EEG preprocessing

Data from 2 participants were deemed too noisy for EEG analyses (final N=18), and the number of sessions per participant varied from 1-4 (mean ± SD: 2.78 ± 0.73). Channel F8-F7 was considered the best (i.e., consistently least noisy) derivative across all data and was used for further analyses. We chose to use the same derivative across all participants rather than individualising it to reduce any variability as a result of channel derivative. Data were preprocessed using the the Oxford Centre for Human Brain Activity (OHBA) Software Library (OSL) (Quinn et al., 2024; van Es et al., 2025). Firstly, data from channel F8-F7 were bandpass filtered between 0.4 Hz and 30 Hz using 5^th^ order Butterworth, downsampled to 100 Hz, and then artifacts were automatically detected using significance levels of p=0.150 for both raw and differenced time series data with segment lengths of both 500 ms and 1000 ms (i.e., four layers of artifact detection). For the main analyses, median values were calculated across the included nights.

##### EEG analyses

Preprocessed data from stages N2 and N3 were used for all further analyses. Spindles were detected using yasa’s *spindles_detect* with 12-15 Hz as the frequency of interest. Where no spindles were detected, these still contributed to the spindle density measure (i.e., number of spindles per minute would be zero), however, these did not contribute towards the spindle amplitude measure as we reasoned that should be dependent on a spindle being present. Considering the at-home data collection procedures, additional data quality checks showed that automated detections had not captured a spindle in one overnight session, i.e., the spindle waxing and waning form. Hence, the data from this session were treated the same as if no spindles were detected (N=1; see supplementary figure including examples of included nights and the excluded night). Slow-oscillations and their coupling with spindles were detected using yasa’s *sw_detect* with the slow oscillation frequency of 0.3-1.5 Hz and spindle frequency of 12-15 Hz.

#### Statistical analyses

Firstly, to explore whether lesion volume relates to sleep measures (SCI and EEG) we used Spearman’s correlations in R Studio (version 2024.12.1). For the actigraphy data, due to the high number of variables of interest, we reduced the dimensionality by using a k-means cluster analysis (R package: *cluster*, function: *cluster_analysis*) using the silhouette method (*fviz_nbclust*) to determine the optimal number of clusters. The actigraphy variables used were: WASO (mins), Sleep Fragmentation Index, Wake Bouts, Sleep Efficiency (%), and Assumed Sleep (mins). Lesion volumes were compared between the two clusters using Welch’s t-test and Cohen’s d effect sizes were computed (R package: *lsr*, function: *cohensD*).

To test whether lesion overlap with the ROIs related to sleep measures (SCI and actigraphy clusters) we explored the data using a random forest approach. Random forest is a machine learning method useful in exploratory data analyses instead of using a confirmatory multiple linear regression approach and can detect interactions and non-linear effects in the data (for further details, see (Fife & D’Onofrio, 2023). We used the *cforest* function within the *party* package in R. The predictor variables were age, sex, lesion volume, and binarised overlaps (yes/no) with reach of the ROIs: amygdala, basal ganglia (accumbens, caudate, pallidum), brainstem, hippocampus, and thalamus.

To explore whether structural disconnection relates to sleep measures (SCI, actigraphy, EEG) we ran voxel-wise non-parametric permutation tests (*randomise* function in FSL (Winkler et al., 2014) with 5000 permutations) controlling for multiple comparisons with threshold-free cluster enhancement (Smith & Nichols, 2009) (tfce), using regression (SCI, EEG) and independent t-test (actigraphy clusters) comparisons.

## Results

Participants included 27 males and 11 females, with a mean (± SD) age of 64 (± 12) years, and mean time since stroke of 95 (± 66) months (see table 1). The mean (± SD) time between the MRI and collection of sleep measures was 22 (± 26) months. The mean lesion volume was 32675 mm^3^ and the distribution of lesions is visualised in figure 1A (note that we did not ‘flip’ any scans to align lesioned hemispheres across patients). The number of participants with a lesion overlapping each of the ROIs is shown in supplementary table S1. The subsample of participants for whom a modified Rankin Scale score was available (N= 26) showed most patients were mild to moderate severity (mRS scores 1=36%, 2=28%, 3=32%).

**Figure 1.**
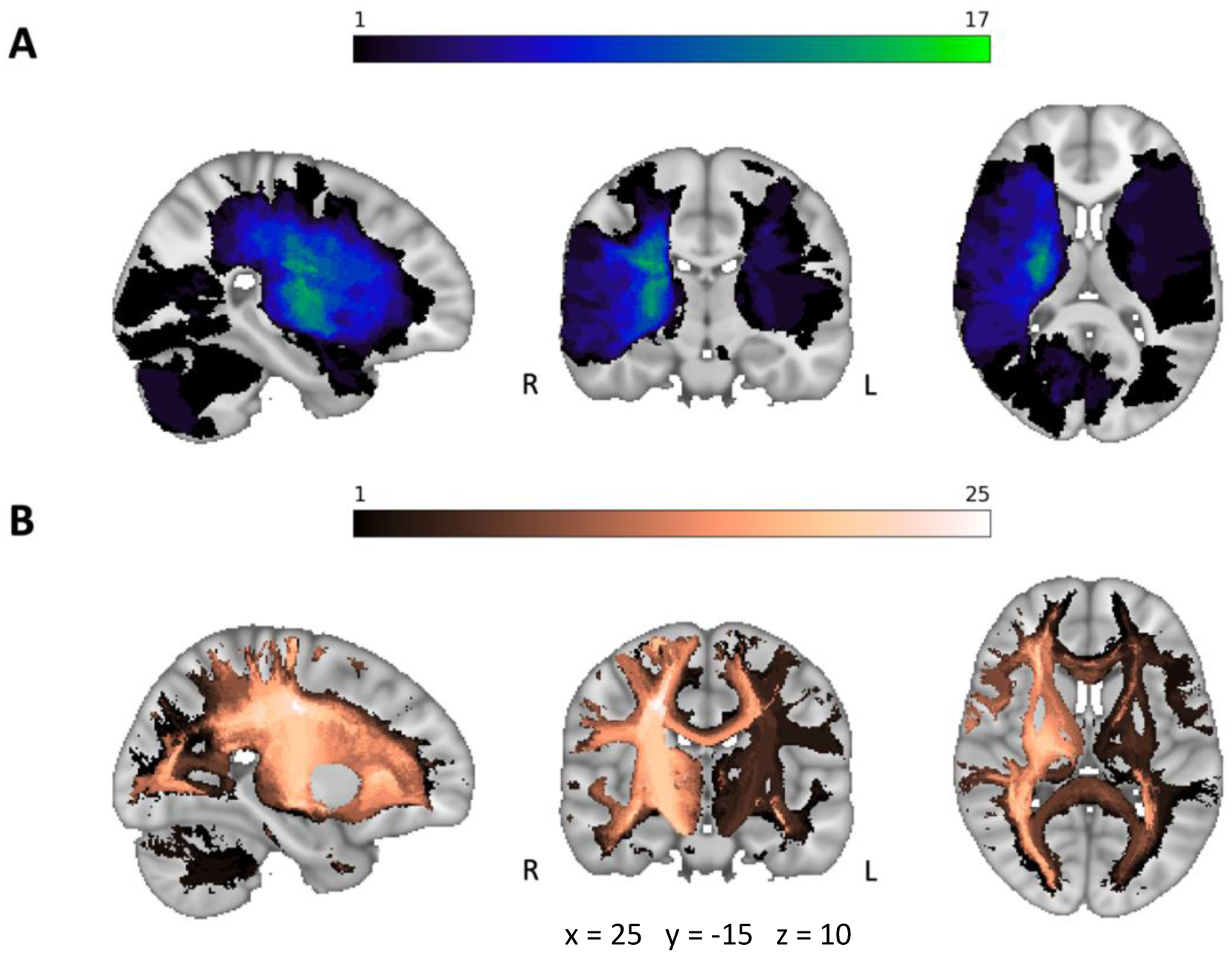
Heatmaps in standard (MNI 1mm) space. **A:** Lesion overlap across all participants (N=38, max overlap N=17). **B:** Structural disconnectome (i.e., voxel-wise white matter estimates disconnected by the stroke lesions) map overlap across all participants (N=38, max overlap N=25). Note that the scans are not ‘flipped’ to align lesioned hemispheres across patients.

### Questionnaires

The completion rate varied across questionnaires and is shown in table 2. Out of the 25 participants who completed the SDSQ, 6 presented with possible narcolepsy, 3 with a possible sleep breathing disorder, 8 with possible restless leg syndrome, 3 with a possible circadian disorder, and none presented with possible parasomnia. Of the subsample who completed the STOP-BANG questionnaire (N=26), 38% were estimated to be at high risk of sleep apnoea (score of 5 or above). Based on the PSQI, 80% of the sample presented with poor subjective sleep (based on a score of 5 points or greater (Buysse et al., 1989)), but only a small proportion (35%) experienced excessive daytime sleepiness (based on a score of 10 or greater (Johns, 1991). A small proportion of the sample reported symptoms suggestive of moderate-severe depression (11%) or anxiety (15%) based on a score of 10 or greater on the PHQ and GAD respectively (Kroenke et al., 2001; Spitzer et al., 2006).

**Table 2.**
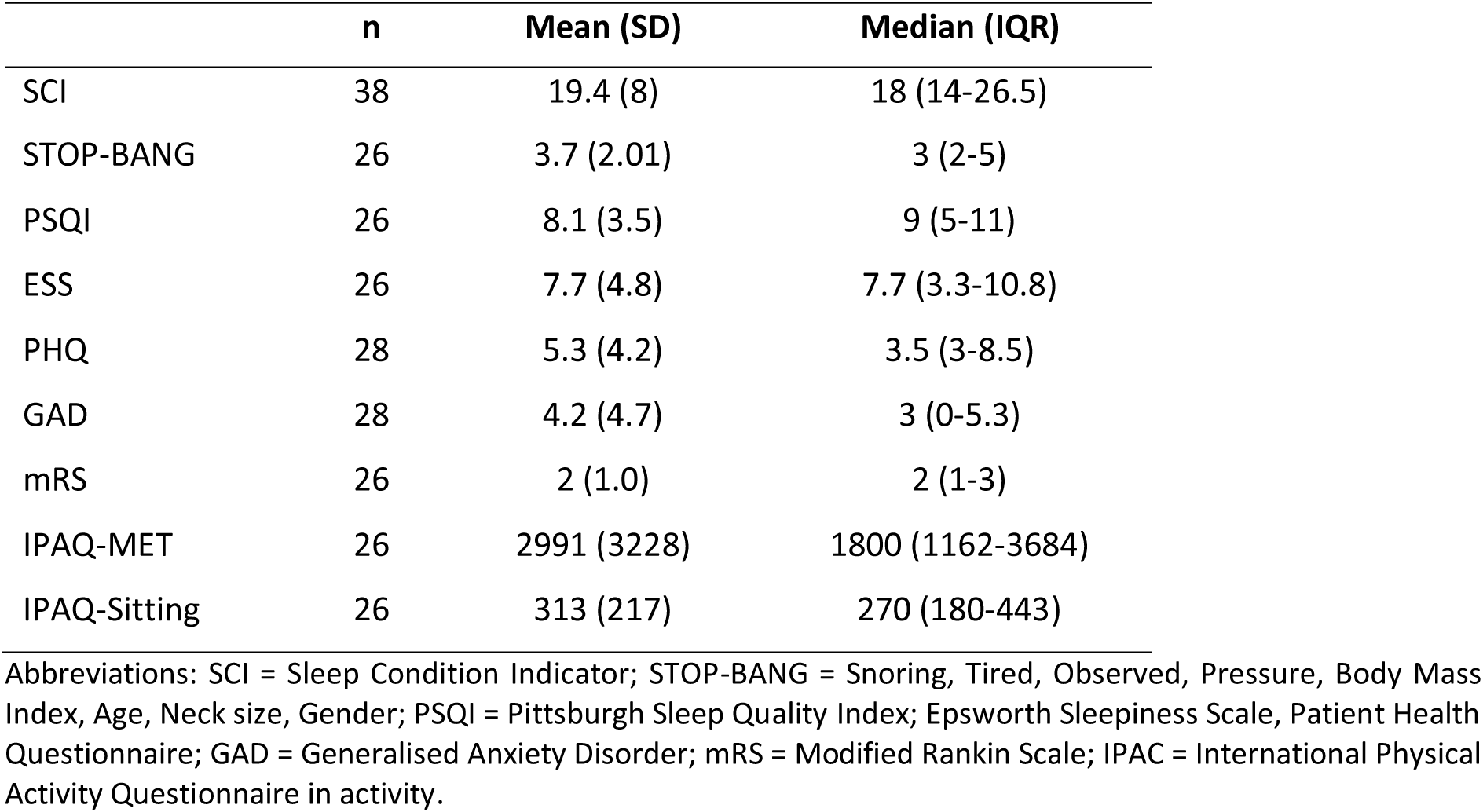
Summary of questionnaires.

The main subjective sleep measure of interest was SCI, a measure of insomnia symptoms, as we have previously found this to be sensitive to differences in sleep between people with acquired brain injury (including stroke) and controls in the hospital and the community (Fleming et al., 2021; Fleming et al., 2020) and the SCI has been validated in stroke (McLaren et al., 2024). The median (interquartile range) SCI score was 18 (14-27) and 21-39% of the sample presented with probable insomnia disorder, based on a score of ≤13 (McLaren et al., 2024) or ≤16 (Espie et al., 2014), respectively. There was no significant correlation between lesion volume and SCI (r_s_=-0.09, p=0.613, see 2A).

We next explored the relationship between lesion overlaps with a priori regions of interest (amygdala, basal ganglia, brainstem, hippocampus, and thalamus) and SCI score. Using a random forest approach, the predictors lesion volume, age, sex, and overlaps with the regions of interest (yes/no) explained only 2% of the variance in SCI score. Variable importance (VI, i.e., root MSE of predicted versus permuted, a higher number indicating more importance) was 1.43 for basal ganglia, 0.31 for amygdala, and no other contributing predictors (VI=0). Based on this, we compared SCI for the variable with the highest importance (basal ganglia ROI), but found no significant differences when comparing SCI score for participants with basal ganglia lesion overlap to those without (t(30.36)=1.61, p=0.119, d = 0.54, see figure 2B).

**Figure 2.**
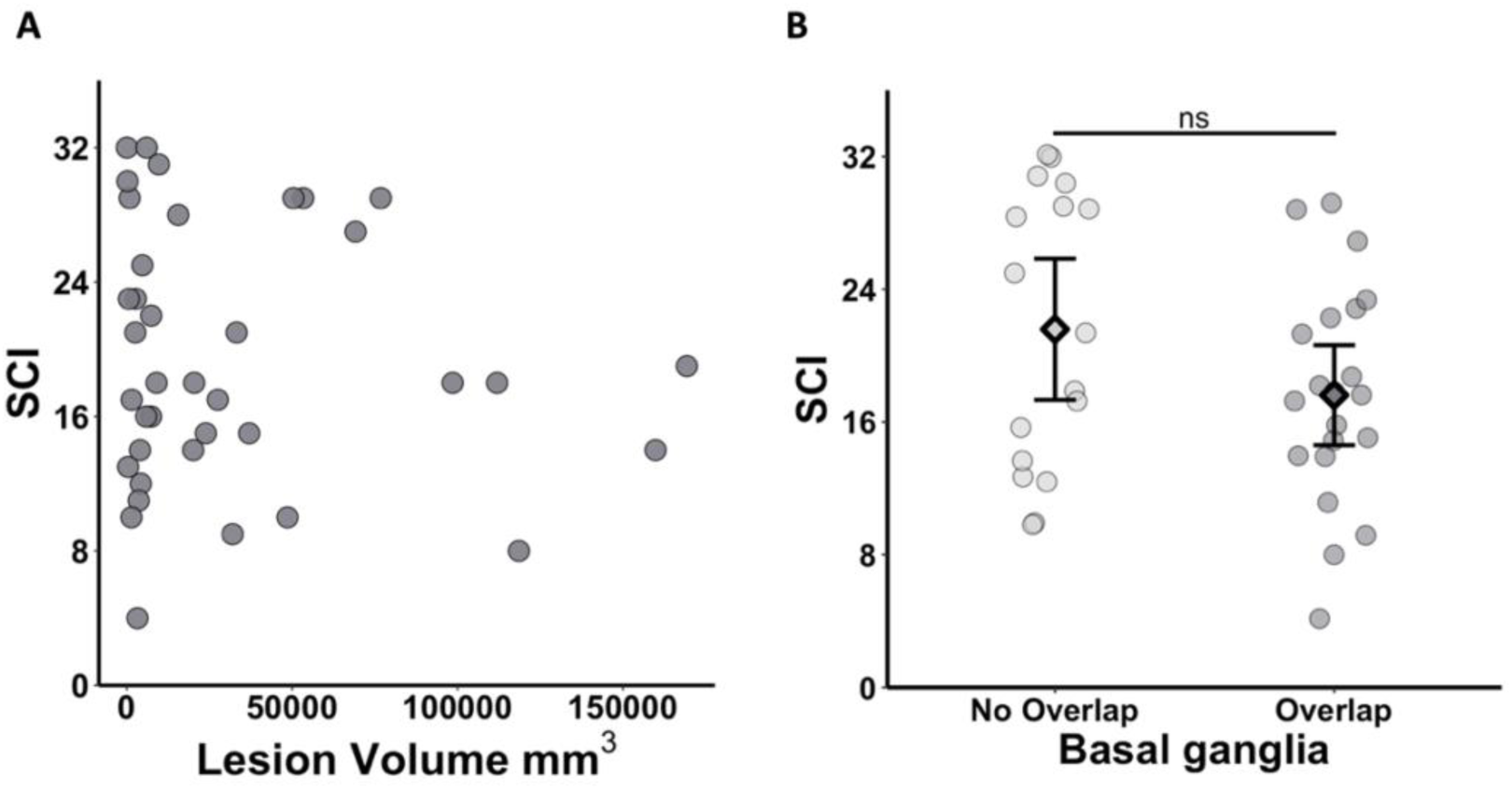
Subjective sleep and lesion characteristics. A: There was no significant relationship between lesion volume and the SCI score (Spearman’s correlation; p > 0.050). **B:** There was no significant difference in SCI score between participants with and without lesions overlapping with the basal ganglia (graph shows mean and standard deviation; p > 0.050). Data are presented as mean (diamond) with error bars reflecting standard error of the mean. A higher SCI score reflects better subjective sleep (fewer symptoms of insomnia).

Finally, we ran a permutation regression analysis between the disconnectome maps and SCI score, but found no significant relationship (p > 0.050, threshold free cluster enhancement (tfce); see data availability section for links to statistical maps).

### Actigraphy

Due to having a high number of actigraphy variables of interest, we used a k-means cluster analysis to reduce the number of comparisons needed. Two clusters (see figure 3) were revealed which we post-hoc labelled “poor” (N=18) and “good” sleepers (N=19). “Poor” sleepers were characterised by more disrupted sleep (longer WASO, higher sleep fragmentation and a greater number of awakenings; see Table 3 for a summary of the actigraphy measures per cluster group). To better understand how these groupings fit into subjective ratings of sleep, we compared the “poor” and “good” sleepers in their SCI score, but no significant differences emerged (t(34.21)=0.16, p=0.873, d = 0.05).

**Figure 3.**
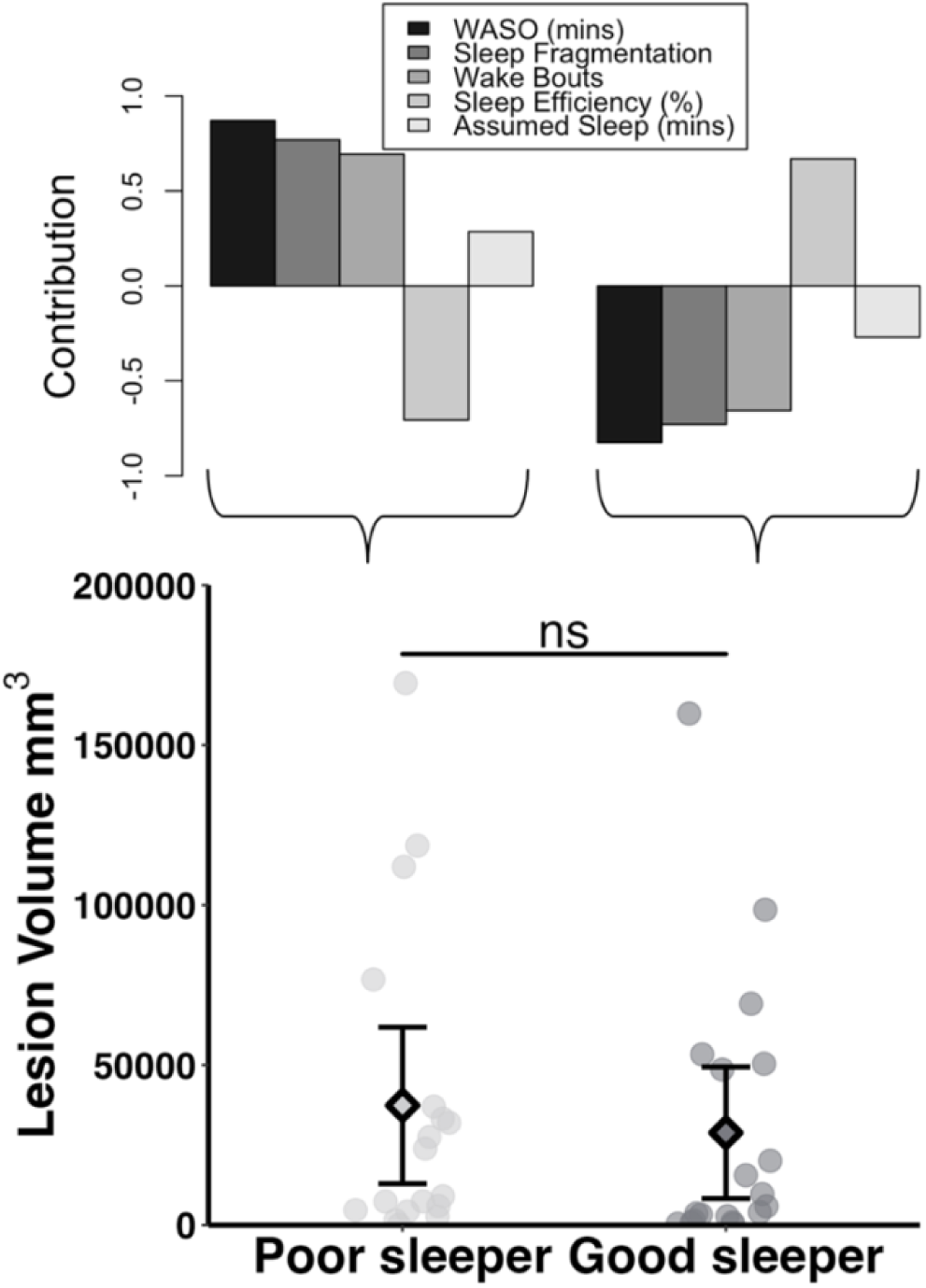
The two groups of sleepers according to the k-means cluster analysis did not show significantly different lesion volume (bottom plot; mean (diamond) and SEM, p > 0.050). The top plot shows the contribution of each sleep measure to each of the clusters.

**Table 3.** Summary of actigraphy sleep variables per cluster group.

| Sleep measure | "Poor" sleeper (N=18) | "Good" sleeper (N=19) |
| --- | --- | --- |
| WASO (mins) | 89.28 ($\pm 18.54$) | 38.89 ($\pm 11.68$) |
| Sleep Fragmentation | 47.59 ($\pm 12.39$) | 24.07 ($\pm 7.96$) |
| Wake Bouts (count) | 51.61 ($\pm 15.67$) | 29.79 ($\pm 6.78$) |
| Sleep Efficiency (%) | 82.86 ($\pm 3.90$) | 91.29 ($\pm 3.21$) |
| Assumed Sleep (mins) | 501.83 ($\pm 71.29$) | 467.16 ($\pm 48.83$) |

Data are shown as mean ( ± standard deviations).

A Welch’s comparison revealed no significant difference in lesion volume between the two types of sleepers (t(33.71)=0.56, p=0.578, d = 0.19; Figure 3). See supplementary figure S3 for scatterplots showing each individual actigraphy variable and lesion volume.

Using a random forest approach, the out-of-bag (i.e., classification) accuracy for “poor” and “good” sleepers was 45.9% with predictors including age, sex, lesion volume and lesion overlaps with the ROIs. None of the ROI overlaps nor lesion volume had a variable importance above 0, with sex and age only just above 0 (sex VI=0.021; age VI=0.004). Thus, the lesion overlaps with ROIs did not seem to aid classification of whether the participants belonged to the poor or good sleeper group.

Finally, using an independent t-test permutation approach, we compared the disconnectome maps for the poor and good sleepers. There were no significant differences (p > 0.050, tfce; see data availability section for links to statistical maps).

### Sleep stages and EEG

In total, 21 of the participants were willing and able to wear the EEG headband at home. However, for two participants it was not possible for them to position the headband in a way to ensure sufficient contact with the scalp over the night to obtain usable data (based on visual inspection of the data by a trained sleep EEG scorer). Furthermore, due to device malfunction, data from one participant was lost. The resulting sleep stage and microstructure measures are shown in table 4. We attempted a k-means cluster analysis, but no clear clusters emerged. Thus, we explored individual relationships for each of the sleep EEG metrics of interest. Due to the exploratory nature, and small sample size, we have not corrected for multiple comparisons and results should be interpreted cautiously.

**Table 4.**
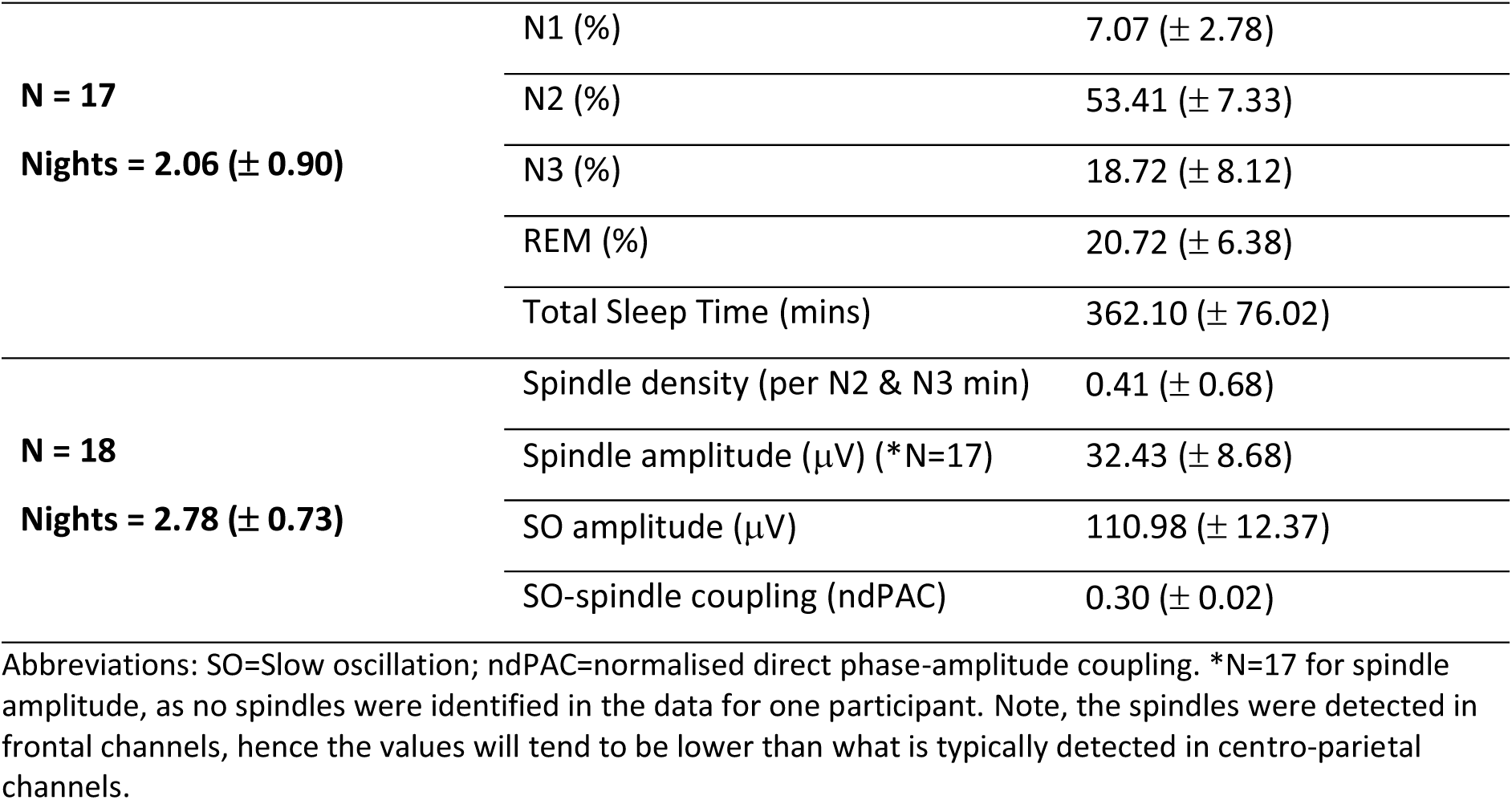
Mean (± standard deviations) of sleep EEG measures.

There were no significant correlations between lesion volume and the percentage of time spent in stages N1 (r_s_=0.20, p=0.432), N2 (r_s_=0.02, p=0.951), N3 (r_s_=0.01, p=0.966), REM (r_s_=-0.20, p=0.438), nor with total sleep time (TST, r_s_=-0.13, p=0.612, all uncorrected, see figure 4). Similarly, there were no significant correlations between lesion volume and spindle density (r_s_=-0.03, p=0.908), spindle amplitude (r_s_=-0.34, p=0.178), SO amplitude (r_s_=-0.22, p=0.384), nor SO-spindle coupling strength (r_s_=-0.04, p=0.882, all uncorrected, see figure 5). Note, for spindle amplitude, data from one participant was removed as no spindles were identified in their data. Results remained unchanged when two outliers were removed (see figures 4 and 5).

**Figure 4.**
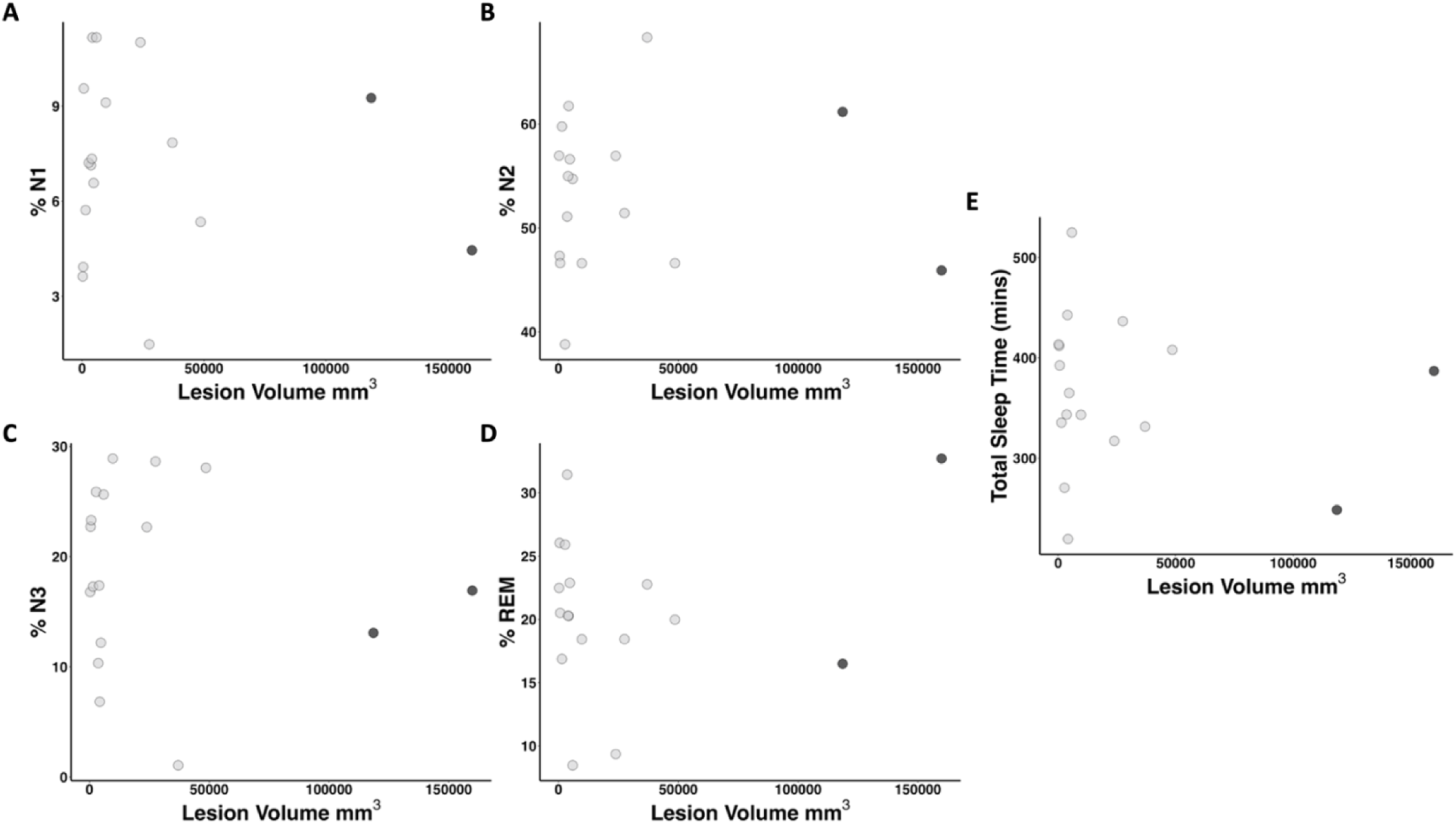
Lesion volume showed no significant relationship with percentage time spent in stages **A:** N1, **B:** N2, **C:** N3, **D:** REM, or with **E:** total sleep time (N=17, all, p > .050). The two outliers are marked in dark grey.

**Figure 5.**
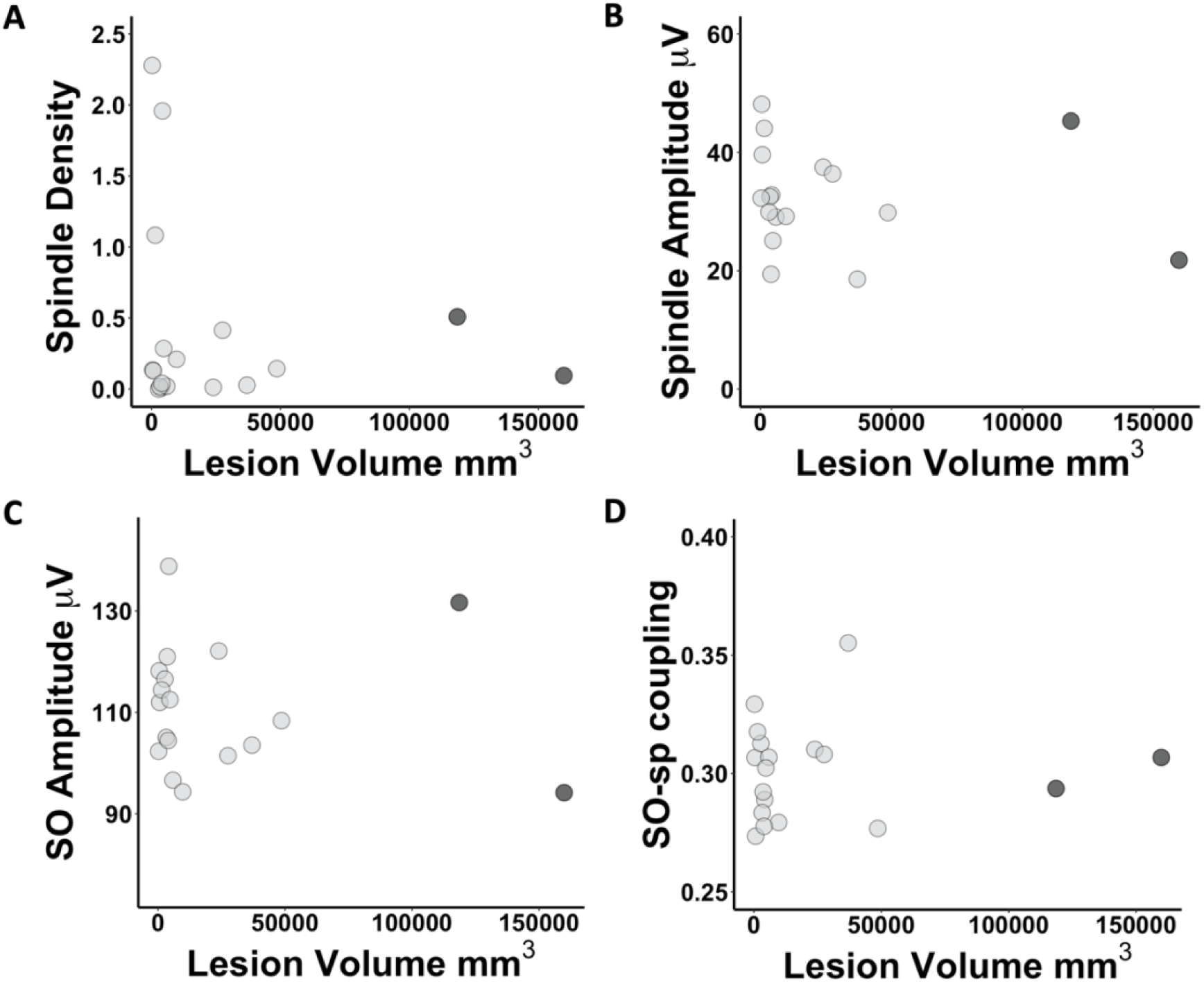
Lesion volume in the subsample (N=18) showed no significant relationship with the following EEG sleep microstructures, **A:** spindle density, **B:** spindle amplitude, **C:** SO amplitude, and **D:** SO-spindle coupling (all p > .050). The two outliers are marked in dark grey.

Considering the small number of participants with useable EEG data who also had lesions overlapping regions of interest (see supplementary table S1), we did not consider it statistically appropriate to perform any comparisons to test for relationships with ROI overlap.

Instead, we ran permutation regression analyses correlating disconnectome maps with spindle density (N=18), spindle amplitude (N=17), SO amplitude (N=18), and SO-spindle coupling (N=18). While there were no significant links with SO amplitude and SO-spindle coupling, there were negative relationships with spindle density and spindle amplitude, i.e., lower spindle density and spindle amplitude were linked to higher probability of disconnection as a result of the stroke lesion shown in figure 6 (p < 0.050, tfce; see data availability section for links to statistical maps).

**Figure 6.**
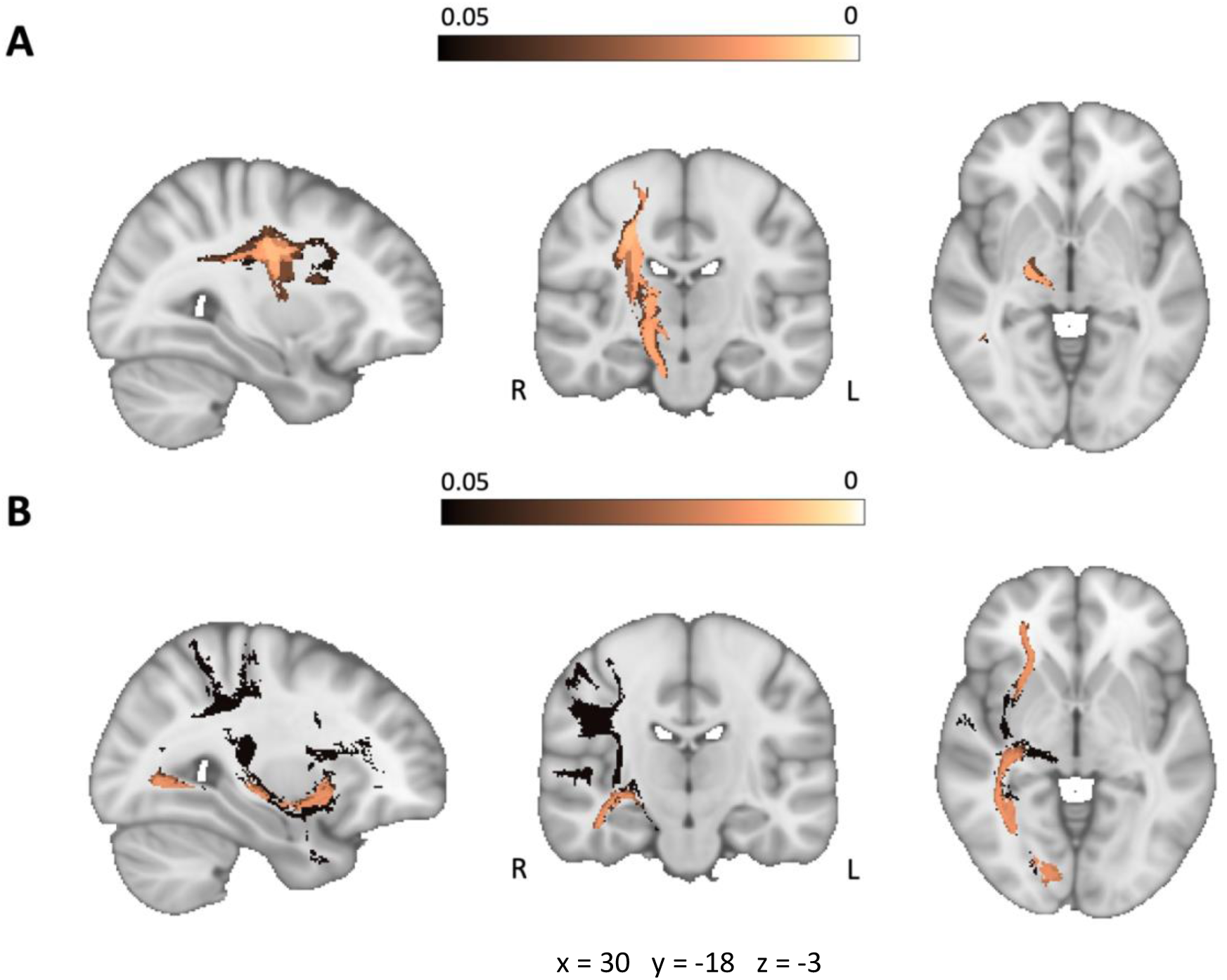
P-value maps from randomise showing a negative relationship between disconnectome maps and **A:** spindle density and **B:** spindle amplitude (p < 0.050, tfce).

## Discussion

In this pragmatic pilot study, we aimed to identify potential relationships between lesion characteristics and sleep outcomes following stroke in chronic stroke survivors, to inform design of future prospective studies. We demonstrated that it is possible to collect an array of sleep variables in a home setting after stroke. However, there was no apparent indication that the lesion size or ROI-based location of stroke lesions relative to brain areas involved in regulation of the sleep-wake cycle related to interindividual variability in self-reported sleep, actigraphy sleep measures, or EEG-based macro architecture variables. In contrast, a data-driven approach revealed that clusters of voxels with a higher probability of structural disconnection voxels were linked to lower spindle density and amplitude. Overall, these results provide preliminary insights into the potential absence or presence of links between stroke lesion characteristics and sleep.

The sample was driven by the existence of a T1-weighted anatomical MRI scan from previous studies in our centre. As such, the participants were typically mild-moderately impaired, there was a wide range in terms of time since stroke, and the group was on average younger than a typical clinical stroke cohort. Participants were not recruited on the basis of having a clinical sleep problem, but the majority were found to have poor sleep quality according to the PSQI and 21-39% presented with probable insomnia disorder (depending on whether a stroke-specific or standard cut-off was used). This is similar (or slightly lower) than the estimated insomnia prevalence based on meta-analysis of stroke studies (Baylan et al., 2020). Additionally, the average values for actigraphy data are broadly similar to previous studies of sleep in chronic community dwelling stroke survivors who typically show more disrupted sleep than controls (Falck et al., 2019; Fleming et al., 2021). Our EEG findings suggest a lower percentage of time spent in N1 sleep and higher percentage in N3 sleep on average than is typically reported (Baglioni et al., 2016), but it is difficult to directly compare our sleep EEG variables to prior studies which are typically done in a lab or hospital setting and early after stroke. However, in comparison to Gottlieb et al. (2021), which was conducted in a home setting, with a comparative sample size (N=28), we also found a slightly lower amount of time spent in N1 sleep (25 vs 48 minutes), more N2 (193 vs 143 minutes) and more N3 (70 vs 66 minutes) accompanied by overall slightly longer total sleep time in our sample (364 vs 320 minutes). It is difficult to speculate as to the reasons underlying these differences, but the study by Gottlieb et al. (2021) restricted their inclusion to ischaemic stroke only, and excluded anyone taking sleep medications such as benzodiazepines or selective serotonin reuptake inhibitors which could have potentially influenced the results found here. Additionally, our average time since stroke was longer and further research is needed to better understand the longitudinal changes in sleep architecture after stroke.

We observed relationships between regions of structural disconnections associated with the stroke lesions and our spindle measures of interest, whereby greater probability of disconnection in white matter regions shown in figure 6 were linked to lower spindle density and amplitude. Reductions in spindle density and amplitude have previously been observed in acute stroke (Bassetti & Aldrich, 2001; Gottselig et al., 2002), with the size of spindle peaks increasing from earliest acute stage to the chronic stage of stroke on the affected side suggesting some level of spindle recovery (Gottselig et al., 2002; Kim et al., 2022). With the present chronic stroke group, we speculate that these spindle characteristics may reflect more permanent reductions in spindle characteristics when stroke lesions disconnect specific structural pathways (although causality cannot be inferred with the present correlational approach). From the disconnectome maps (figure 6), the observed effects were lateralised to the right hemisphere, however, this would likely be reflective of the sample who mostly had right-hemispheric lesions. From visual characterisation of the clusters, both appear to overlap with right superior longitudinal fasciculus, arcurate segment, and cortico-spinal tracts, with the spindle density disconnection cluster particularly overlapping with the right pons, fronto-striatal, and anterior thalamic projections, whilst the spindle amplitude disconnection map tending to overlap with right anterior commissure, optic radiations, fornix, and inferior fronto-occipital fasciculus. These maps of structural disconnection can help guide future confirmatory studies characterising lesion and spindle features after stroke. Of note is the relationship between the thalamic projection disconnection and spindle density which aligns with the idea that spindles are generated in thalamus and projected to cortical regions via thalamo-cortical pathways (Piantoni et al., 2016; Steriade, 2006). On the other hand, spindle amplitude disconnection appears more widespread, and although we did not find any links between stroke lesion size and spindle amplitude, this has been reported previously in acute stroke (Gottselig et al., 2002). Taken together, we speculate that spindle density disturbance might be linked to more specific thalamic pathway lesions, whereas spindle amplitude reductions may reflect more non-specific disruption.

Whilst we were able to collect a variety of sleep outcome measures in this population, the collection of sleep EEG data in particular, was challenging. Participants were generally willing and able to wear the low-density commercial EEG headband in their homes, but data quality varied and required extensive reviewing and rescoring by trained sleep scorers, rather than relying on the automated outputs from the commercial system. This resulted in fewer participants with available data than we had aimed for, and consequently all analyses should be interpreted cautiously. We were able to quantify oscillatory activity during sleep, including slow wave and sleep spindle characteristics, but were unable to determine whether these characteristics differed between the ipsilesional and contralesional hemisphere. A previous study found no significant differences in the time spent in each stage of sleep when comparing the ipsilesional and contralesional hemispheres in chronic stroke survivors (Gottlieb et al., 2021), but since the characteristics of slow oscillations and spindles vary across regions in the brain (Nir et al., 2011), these microarchitecture measures may be more sensitive to underlying tissue damage than sleep stage variables.

Due to the low-density EEG system used, we were unable to test for sleep apnoea directly. Previous research has suggested that even when patients do not report sleep apnoea, a high proportion do present with moderate to severe obstructive sleep apnoea on the basis of the apnoea-hypopnea index (AHI) (Gottlieb et al., 2021). In their study, controlling for AHI attenuated the difference in sleep architecture characteristics between patients and controls, but the effect size of the differences remained similar, suggesting that sleep apnoea alone is not responsible for the relative increase in lighter (N1/N2) and decrease in slow wave sleep (N3) compared to controls without stroke.

A key limitation of this study is the small sample size. Although we acknowledge that it is difficult to draw conclusions based on this sample, the findings reported here could be representative of sleep changes after stroke in larger, more heterogenous cohorts; that lesion location and extent may not predict sleep problems and instead other demographic and clinical factors could be more important long-term. For example, factors such as post-stroke pain, depression, anxiety, inactivity, and co-morbidities such as diabetes and dementia could all influence sleep quality and architecture (Hwang & Kim, 2023). That being said, our data-driven analysis approach highlights subtle results regarding structural disconnection and spindle characteristics that require further investigation with a more robust sample size.

In conclusion, this study is an initial proof-of-principle that it is possible to utilise a multimodal approach to measure the impact of stroke lesions on sleep measures. Although we were unable to identify any candidate regions to pursue in future studies investigating subjective and actigraphy-based measures of sleep, this study adds to a growing body of literature highlighting the complex relationship between sleep EEG characteristics and stroke. Future research is needed to better understand the predictors of sleep problems, to guide treatment development and improve rehabilitation outcomes long term.

## Data availability

T-statistics maps are available at Neurovault via the following link: https://identifiers.org/neurovault.collection:20424. The study preregistration can be found here: https://osf.io/khqn8. Data derivatives and analysis scripts will be made available here: https://osf.io/vbu7c/. Raw data are available upon reasonable request from the corresponding author.

## Acknowledgements

We would like to thank the participants for taking part in this study. Thanks to Tom Smejka, Ellie Macey, Zeena-Britt Sanders and Marilien Marzolla who were involved in collection of some of the MR and sleep data included here.

## Author Contributions

**Conceptualization:** AáVG, MFK

**Methodology:** AáVG, MW, MFK

**Data curation:** AáVG, MW, BR, KS, PLW, OP

**Formal analysis:** AáVG

**Investigation:** MW, BR

**Writing – Original Draft:** AáVG, MFK

**Writing – Review & Editing:** AáVG, MW, BR, KS, PLW, HEW, HB, CJS, IFG, OP, HJB, MKF

**Resources:** HEW, HB, CJS, IFG, HJB, MKF

**Supervision:** MKF

**Funding acquisition:** MKF, HJB

## Rights Retention

This research is funded in whole, or in part, by the Wellcome Trust [222446/Z/21/Z, 203139/Z/16/Z and 203139/A/16/Z]. For the purpose of open access, the author has applied a CCBY public copyright license to any Author Accepted Manuscript version arising from this submission.

## References

Arnal, P. J., Thorey, V., Debellemaniere, E., Ballard, M. E., Bou Hernandez, A., Guillot, A., Jourde, H., Harris, M., Guillard, M., & Van Beers, P. (2020). The Dreem Headband compared to polysomnography for electroencephalographic signal acquisition and sleep staging. Sleep, 43(11), zsaa097. 10.1093/sleep/zsaa097

Baglioni, C., Nissen, C., Schweinoch, A., Riemann, D., Spiegelhalder, K., Berger, M., Weiller, C., & Sterr, A. (2016). Polysomnographic characteristics of sleep in stroke: a systematic review and meta-analysis. PLoS One, 11(3), e0148496. 10.1371/journal.pone.0148496

Baillieul, S., Denis, C., Barateau, L., Arquizan, C., Detante, O., Pépin, J.-L., Dauvilliers, Y., & Tamisier, R. (2023). The multifaceted aspects of sleep and sleep-wake disorders following stroke. Revue Neurologique, 179(7), 782–792. 10.1016/j.neurol.2023.08.004

Bassetti, C. L., & Aldrich, M. S. (2001). Sleep electroencephalogram changes in acute hemispheric stroke. Sleep medicine, 2(3), 185–194. 10.1016/S1389-9457(00)00071-X

Baylan, S., Griffiths, S., Grant, N., Broomfield, N. M., Evans, J. J., & Gardani, M. (2020). Incidence and prevalence of post-stroke insomnia: a systematic review and meta-analysis. Sleep Medicine Reviews, 49, 101222. 10.1016/j.smrv.2019.101222

Buysse, D. J., Reynolds, C. F., 3rd, Monk, T. H., Berman, S. R., & Kupfer, D. J. (1989). The Pittsburgh Sleep Quality Index: a new instrument for psychiatric practice and research. Psychiatry Res., 28(2), 193–213. 10.1016/0165-1781(89)90047-4

Cai, H., Wang, X.-P., & Yang, G.-Y. (2021). Sleep disorders in stroke: an update on management. Aging and disease, 12(2), 570. 10.14336/AD.2020.0707

De Schotten, M. T., Bizzi, A., Dell’Acqua, F., Allin, M., Walshe, M., Murray, R., Williams, S. C., Murphy, D. G., & Catani, M. (2011). Atlasing location, asymmetry and inter-subject variability of white matter tracts in the human brain with MR diffusion tractography. Neuroimage, 54(1), 49–59. 10.1016/j.neuroimage.2010.07.055

Diekelmann, S., & Born, J. (2010). The memory function of sleep. Nat. Rev. Neurosci., 11(2), 114–126. 10.1038/nrn2762

Duss, S. B., Seiler, A., Schmidt, M. H., Pace, M., Adamantidis, A., Müri, R. M., & Bassetti, C. L. (2017). The role of sleep in recovery following ischemic stroke: a review of human and animal data. Neurobiology of sleep and circadian rhythms, 2, 94–105. 10.1016/j.nbscr.2016.11.003

Espie, C. A., Kyle, S. D., Hames, P., Gardani, M., Fleming, L., & Cape, J. (2014). The Sleep Condition Indicator: a clinical screening tool to evaluate insomnia disorder. BMJ Open, 4(3), e004183. 10.1136/bmjopen-2013-004183

Falck, R. S., Best, J. R., Davis, J. C., Eng, J. J., Middleton, L. E., Hall, P. A., & Liu-Ambrose, T. (2019). Sleep and cognitive function in chronic stroke: a comparative cross-sectional study. Sleep, 42(5), zsz040. 10.1093/sleep/zsz040

Fife, D. A., & D’Onofrio, J. (2023). Common, uncommon, and novel applications of random forest in psychological research. Behavior research methods, 55(5), 2447–2466. 10.3758/s13428-022-01901-9

Fleming, M. K., Smejka, T., Henderson Slater, D., Chiu, E. G., Demeyere, N., & Johansen-Berg, H. (2021). Self-reported and objective sleep measures in stroke survivors with incomplete motor recovery at the chronic stage. Neurorehabilitation and Neural Repair, 35(10), 851–860. 10.1177/15459683211029889

Fleming, M. K., Smejka, T., Henderson Slater, D., van Gils, V., Garratt, E., Yilmaz Kara, E., & Johansen-Berg, H. (2020). Sleep disruption after brain injury is associated with worse motor outcomes and slower functional recovery. Neurorehabilitation and Neural Repair, 34(7), 661–671. 10.1177/1545968320929669

Foulon, C., Cerliani, L., Kinkingnehun, S., Levy, R., Rosso, C., Urbanski, M., Volle, E., & Thiebaut de Schotten, M. (2018). Advanced lesion symptom mapping analyses and implementation as BCBtoolkit. Gigascience, 7(3), giy004. 10.1093/gigascience/giy004

Fulk, G. D., Boyne, P., Hauger, M., Ghosh, R., Romano, S., Thomas, J., Slutzky, A., & Klingman, K. (2020). The impact of sleep disorders on functional recovery and participation following stroke: a systematic review and meta-analysis. Neurorehabilitation and Neural Repair, 34(11), 1050–1061. 10.1177/1545968320962501

Gottlieb, E., Egorova, N., Khlif, M. S., Khan, W., Werden, E., Pase, M. P., Howard, M., & Brodtmann, A. (2020). Regional neurodegeneration correlates with sleep–wake dysfunction after stroke. Sleep, 43(9), zsaa054. 10.1093/sleep/zsaa054

Gottlieb, E., Khlif, M. S., Bird, L., Werden, E., Churchward, T., Pase, M. P., Egorova, N., Howard, M. E., & Brodtmann, A. (2021). Sleep architectural dysfunction and undiagnosed obstructive sleep apnea after chronic ischemic stroke. Sleep medicine, 83, 45–53. 10.1016/j.sleep.2021.04.011

Gottlieb, E., Landau, E., Baxter, H., Werden, E., Howard, M. E., & Brodtmann, A. (2019). The bidirectional impact of sleep and circadian rhythm dysfunction in human ischaemic stroke: a systematic review. Sleep Medicine Reviews, 45, 54–69. 10.1016/j.smrv.2019.03.003

Gottselig, J., Bassetti, C., & Achermann, P. (2002). Power and coherence of sleep spindle frequency activity following hemispheric stroke. Brain, 125(2), 373–383. 10.1093/brain/awf021

Hasan, F., Gordon, C., Wu, D., Huang, H.-C., Yuliana, L. T., Susatia, B., Marta, O. F. D., & Chiu, H.-Y. (2021). Dynamic prevalence of sleep disorders following stroke or transient ischemic attack: systematic review and meta-analysis. Stroke, 52(2), 655–663. 10.1161/STROKEAHA.120.029847

Hwang, H.-S., & Kim, H. (2023). Factors affecting the quality of sleep and social participation of stroke patients. Brain Sciences, 13(7), 1068. 10.3390/brainsci13071068

Iber. (2007). The AASM Manual for the Scoring of Sleep and Associated Events : Rules. Terminology and Technical Specification. https://ci.nii.ac.jp/naid/10024500923/

Jenkinson, M., Beckmann, C. F., Behrens, T. E., Woolrich, M. W., & Smith, S. M. (2012). Fsl. Neuroimage, 62(2), 782–790. 10.1016/j.neuroimage.2011.09.015

Johns, M. W. (1991). A new method for measuring daytime sleepiness: the Epworth sleepiness scale. Sleep, 14(6), 540–545. 10.1093/sleep/14.6.540

Kim, J., Guo, L., Hishinuma, A., Lemke, S., Ramanathan, D. S., Won, S. J., & Ganguly, K. (2022). Recovery of consolidation after sleep following stroke—interaction of slow waves, spindles, and GABA. Cell reports, 38(9), 110426. 10.1016/j.celrep.2022.110426

Kroenke, K., Spitzer, R., Williams, J., & Dsw, B. (2001). Validity of a brief depression severity measure. Journal of General Internal Medicine, 16, 1525–1497.2001. 10.1046/j.1525-1497.2001.016009606.x

Liu, L., Wang, W., Gao, N., Jia, T., Guo, L., Geng, L., & Ma, Y. (2023). Risk factors of disturbed sleep phases to posterior circulation cerebral infarctions: A single-center retrospective study. Medicine, 102(41), e35479. 10.1097/MD.0000000000035479

Lutkenhoff, E. S., Rosenberg, M., Chiang, J., Zhang, K., Pickard, J. D., Owen, A. M., & Monti, M. M. (2014). Optimized brain extraction for pathological brains (optiBET). PLoS One, 9(12), e115551. 10.1371/journal.pone.0115551

McLaren, D. M., Evans, J., Baylan, S., Harvey, M., Montgomery, M. C., & Gardani, M. (2024). Assessing insomnia after stroke: a diagnostic validation of the Sleep Condition Indicator in self-reported stroke survivors. BMJ Neurology Open, 6(2), e000768. 10.1136/bmjno-2024-000768

Mekky, J., Hafez, N., Kholy, O. E., Elsalamawy, D., & Gaber, D. (2023). Impact of site, size and severity of ischemic cerebrovascular stroke on sleep in a sample of Egyptian patients a polysomnographic study. BMC neurology, 23(1), 387. 10.1186/s12883-023-03438-6

Nir, Y., Staba, R. J., Andrillon, T., Vyazovskiy, V. V., Cirelli, C., Fried, I., & Tononi, G. (2011). Regional slow waves and spindles in human sleep. Neuron, 70(1), 153–169. 10.1016/j.neuron.2011.02.043

Piantoni, G., Halgren, E., & Cash, S. S. (2016). The contribution of thalamocortical core and matrix pathways to sleep spindles. Neural plasticity, 2016(1), 3024342. 10.1155/2016/3024342

Quinn, A., van Es, M., Gohil, C., & Woolrich, M. (2024). The osl-ephys Python package for the analysis of electrophysiological data. Zenodo. 10.5281/zenodo.14008696

Smith, S. M., & Nichols, T. E. (2009). Threshold-free cluster enhancement: addressing problems of smoothing, threshold dependence and localisation in cluster inference. Neuroimage, 44(1), 83–98. 10.1016/j.neuroimage.2008.03.061

Spitzer, R. L., Kroenke, K., Williams, J. B., & Löwe, B. (2006). A brief measure for assessing generalized anxiety disorder: the GAD-7. Archives of internal medicine, 166(10), 1092–1097. 10.1001/archinte.166.10.1092

Steriade, M. (2006). Grouping of brain rhythms in corticothalamic systems. Neuroscience, 137(4), 1087–1106. 10.1016/j.neuroscience.2005.10.029

Thiebaut de Schotten, M., Dell’Acqua, F., Ratiu, P., Leslie, A., Howells, H., Cabanis, E., Iba-Zizen, M., Plaisant, O., Simmons, A., & Dronkers, N. (2015). From Phineas Gage and Monsieur Leborgne to HM: revisiting disconnection syndromes. Cerebral Cortex, 25(12), 4812–4827. 10.1093/cercor/bhv173

van Es, M. W., Gohil, C., Quinn, A. J., & Woolrich, M. W. (2025). osl-ephys: A Python toolbox for the analysis of electrophysiology data. Frontiers in neuroscience, 19, 1522675.

Wang, R., Benner, T., Sorensen, A. G., & Wedeen, V. J. (2007). Diffusion toolkit: a software package for diffusion imaging data processing and tractography. Proc Intl Soc Mag Reson Med,

Wesselius, H. M., Van Den Ende, E. S., Alsma, J., Ter Maaten, J. C., Schuit, S. C., Stassen, P. M., de Vries, O. J., Kaasjager, K. H., Haak, H. R., & Van Doormaal, F. F. (2018). Quality and quantity of sleep and factors associated with sleep disturbance in hospitalized patients. JAMA internal medicine, 178(9), 1201–1208. 10.1001/jamainternmed.2018.2669

Winkler, A. M., Ridgway, G. R., Webster, M. A., Smith, S. M., & Nichols, T. E. (2014). Permutation inference for the general linear model. Neuroimage, 92, 381–397. 10.1016/j.neuroimage.2014.01.060

