## Supplementary materials for "Exploring the relationship between stroke lesion characteristics and sleep in chronic stroke survivors"

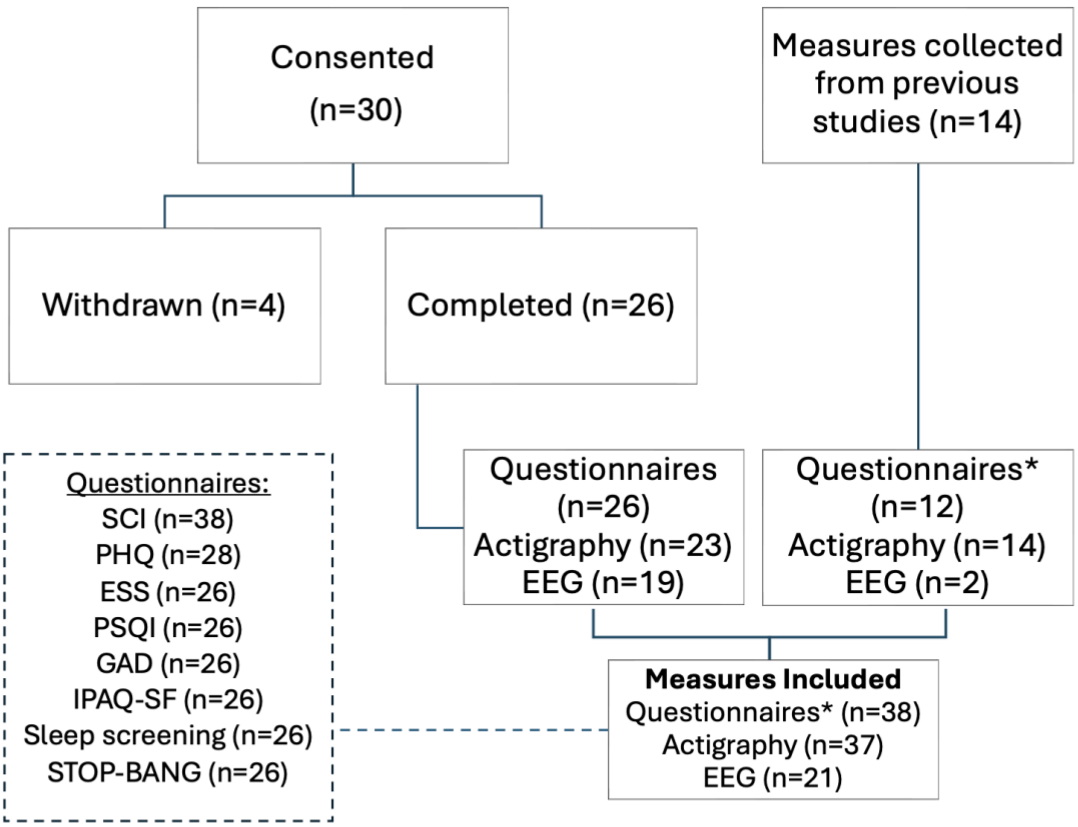


**Figure S1.** Data availability diagram.
SCI: Sleep Condition Indicator, ESS: Epworth Sleepiness Scale, PSQI: Pittsburgh Sleep Quality Index, PHQ: Patient Health Questionnaire, GAD: Generalised Anxiety Disorder, IPAQ-SF: International Physical Activity Questionnaire-Short Form. *Questionnaires – n refers to SCI as minimum.

**Table S1.** Number of participants for each sleep measure with a lesion overlapping the regions of interest.

| | **SCI (max N=38)** | **Actigraphy (max N=37)** | **EEG (max N=18)** |
|---|---|---|---|
| Brainstem | 2 | 2 | 1 |
| Accumbens | 0 | 0 | 0 |
| Amygdala | 8 | 7 | 2 |
| Caudate | 20 | 19 | 8 |
| Hippocampus | 4 | 4 | 2 |
| Pallidum | 16 | 15 | 5 |
| Thalamus | 15 | 14 | 4 |

Abbreviations: SCI=Sleep Condition Indicator; EEG=electroencephalography.

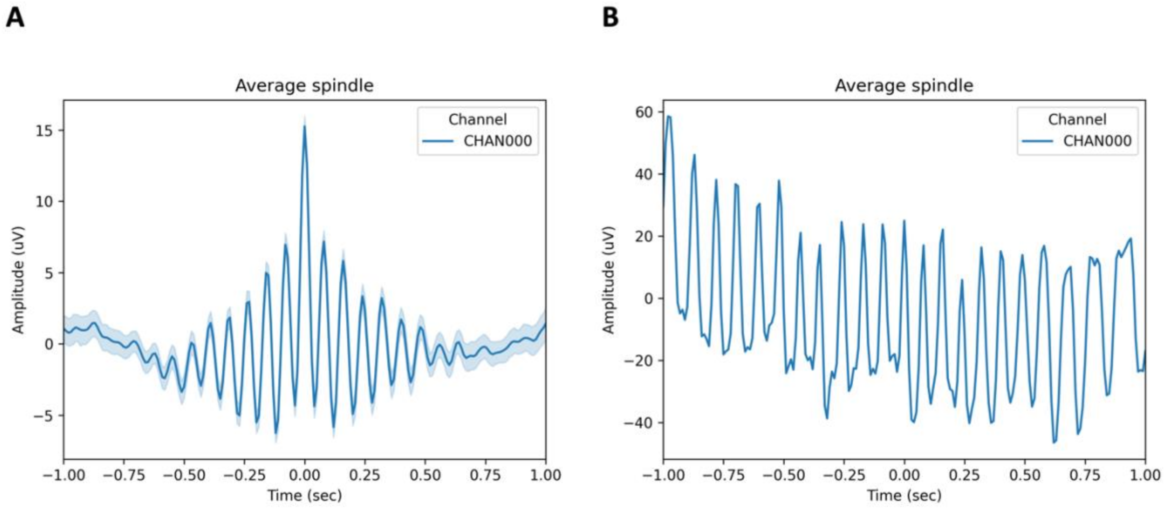


**Figure S2.** Examples from manual spindle data quality check showing **A:** average across multiple spindles and **B:** noise with only one identified spindle which was removed from further analyses.

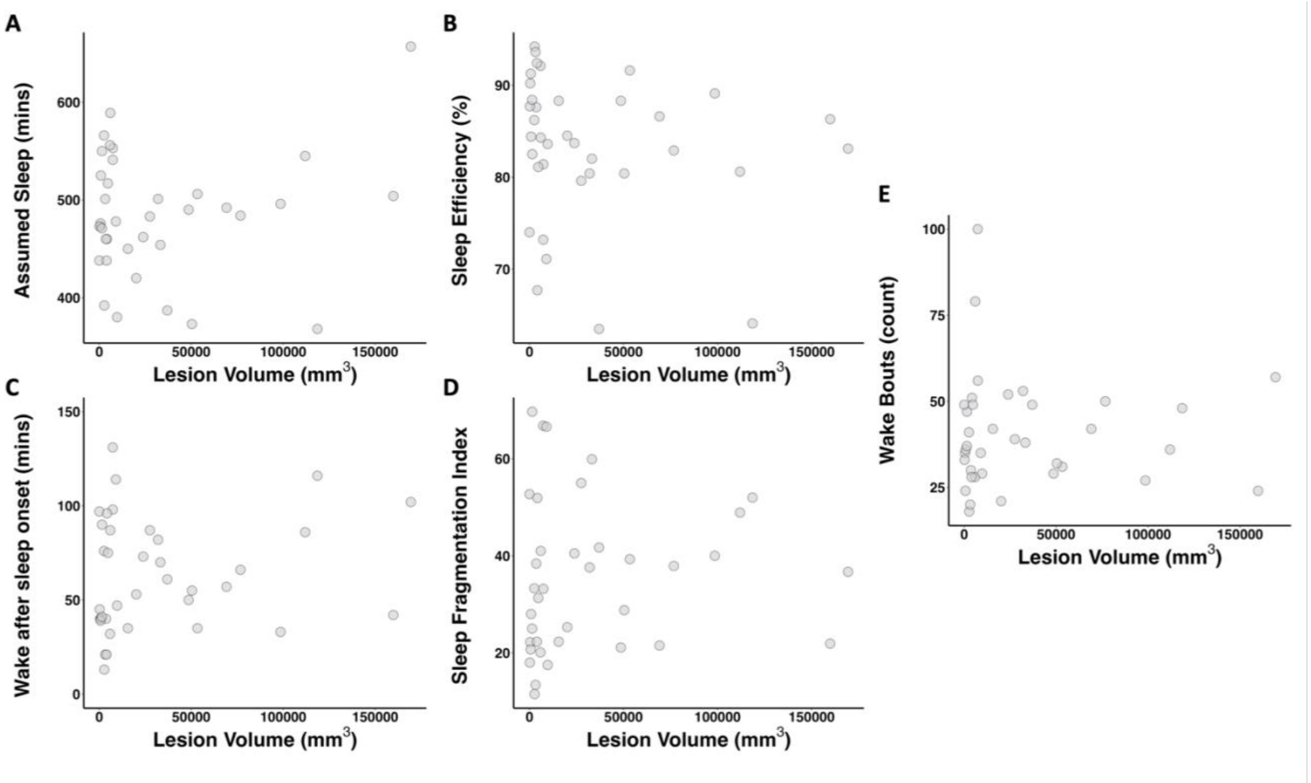


**Figure S3.** Scatterplots of lesion volume with actigraphy variables **A:** Assumed sleep (minutes), **B:** Sleep Efficiency (%), **C:** Wake after sleep onset (WASO; min), **D:** Sleep fragmentation, **E:** Wake bouts (count). There were no signification correlations.

**Table S2. Mean (± standard deviations) of sleep stage measures, time in minutes**

| | | |
|---|---|---|
| **N = 17**<br>**Nights = 2.06 (± 0.90)** | N1 | 25.2 (± 11.4) |
| | N2 | 193.0 (± 43.9) |
| | N3 | 70.1 (± 36.9) |
| | REM | 75.4 (± 26.4) |
| | Total Sleep Time (mins) | 364.22 (± 76.35) |
